# Improvements in cardiovascular health over the perinatal period predicts lower postpartum psychological distress

**DOI:** 10.1101/2023.12.22.23300475

**Authors:** Shannon D. Donofry, Riley J. Jouppi, Christine C. Call, Rachel P. Kolko Conlon, Michele D. Levine

## Abstract

**Background:** Adverse cardiovascular events during pregnancy (e.g., pre-eclampsia) occur at higher rates among individuals with pre-pregnancy overweight or obesity (body mass index [BMI]≥25kg/m^2^) and have been associated with postpartum depression. However, it is unclear whether cardiovascular health (CVH), defined more holistically than the absence of cardiovascular conditions in pregnancy, relates to postpartum psychological functioning. The present study examined whether changes in CVH during the perinatal period predicted postpartum psychological functioning among individuals with pre-pregnancy BMI≥25kg/m^2^.

**Methods:** Individuals (N=226; Mage=28.43±5.4 years; MBMI=34.17±7.15kg/m^2^) were recruited when their pregnancies were 12-20 weeks gestation (M=15.64±2.45 weeks) for a longitudinal study of health and well-being. Participants completed the Center for Epidemiological Studies Depression Scale (CES-D) and Perceived Stress Scale (PSS) and reported on CVH behaviors (dietary intake, physical activity, nicotine exposure, and sleep) at baseline and at 6-months postpartum. BMI and CVH behaviors were coded according to the American Heart Association’s Life’s Essential 8 to create a CVH score at both timepoints. Linear regression analyses were performed to examine whether change in CVH related to postpartum CES-D and PSS scores. Because sleep was only measured in a subset of participants (n=114), analyses were conducted with and without sleep included. Baseline CVH, CES-D and PSS scores, and demographic factors were included as covariates in all models.

**Results:** Improved CVH was associated with lower postpartum CES-D (*β*=-0.18, *p*<0.01) and PSS (*β*=-0.13, *p*=0.02) scores when excluding sleep. Compared to those whose CVH improved by >1SD from pregnancy to 6-months postpartum, individuals whose CVH worsened by >1SD scored 6.42 points higher on the CESD (*M*CESD=15.25±10.92 vs. 8.52±6.90) and 6.12 points higher on the PSS (*M*PSS=24.45±8.29 vs. 17.83±8.70). However, when including sleep, these relationships were no longer significant (*p*s>0.4).

**Conclusions:** Improvements in CVH from early pregnancy to 6-months postpartum were associated with lower postpartum depressive symptoms and perceived stress. However, these relationships were no longer significant when including sleep in the CVH metric, potentially due to the large reduction in sample size. These data suggest that intervening during pregnancy to promote CVH may improve postpartum psychological functioning among high-risk individuals.

## Introduction

Pregnancy is a period in which significant physiologic changes occur in the cardiovascular system to meet the additional metabolic demands associated with maintaining fetal growth and development ^1^. Difficulty adapting to these demands is associated with adverse pregnancy outcomes such as gestational hypertension, preeclampsia and eclampsia, and gestational diabetes ^1^, which occur in about 33% of individuals, are the primary cause of maternal mortality in the United States ^2,3^, and predict future cardiovascular disease (CVD) risk, even among individuals for whom these conditions resolve following delivery ^4,5^. Given that individuals engage with the healthcare system frequently during pregnancy, pregnancy represents not only a period of heightened vulnerability for CVD but also a window of opportunity to positively influence the health trajectories of birthing individuals.

In addition to being at elevated risk for CVD later in life, individuals who are diagnosed with pregnancy-related cardiovascular conditions are more likely to experience postpartum psychological distress, particularly symptoms of depression. A recent meta-analysis of 13 studies demonstrated that hypertensive disorders of pregnancy were associated with higher severity of self-reported postpartum depressive symptoms ^6^. Further, data from a large epidemiological study of nearly 5,000 individuals found evidence that those diagnosed with preeclampsia were more than twice as likely to develop postpartum depression compared to those with normotensive pregnancies ^7^. Gestational diabetes has also been linked to higher risk of postpartum depression. Two recent meta-analyses covering data from ∼4 million individuals with limited overlap in study inclusion found evidence that the odds of postpartum depression were 32-59% higher among individuals whose pregnancies were affected by gestational diabetes compared to those whose were not ^8,9^. Comparatively little research has been conducted to examine the effect of cardiovascular conditions in pregnancy on other postpartum psychological outcomes, despite the fact that symptoms such as anxiety, obsessions, compulsions, and mania are relatively common during the postpartum period ^10,11^. However, one study leveraging data from a nationwide health registry in Denmark demonstrated that cardiovascular conditions in pregnancy such as gestational hypertension and diabetes were associated with higher risk not only of depression, but also symptoms of psychosis and acute stress ^12^. It is important to note that psychiatric symptoms during the perinatal period have a detrimental impact on the health and well-being of the birthing person, as well as infant and child outcomes ^13–17^. Taken together, these data suggest that interventions targeting cardiovascular risk during pregnancy may improve maternal psychological functioning, which may in turn exert downstream benefits on child health and development.

Although previous research has established a link between pregnancy-related cardiovascular conditions and risk for depression during the postpartum period, few studies have explored how indicators of cardiovascular health (CVH) more broadly conceptualized than solely the presence or absence of a CVD diagnosis during pregnancy affects postpartum psychological functioning. To more effectively identify individuals at risk for CVD before the onset of diagnosable disease, the American Heart Association (AHA) developed the Life’s Simple 7 CVH framework, yielding a composite indicator of CVH that encompasses the seven behavioral and physiological factors most closely associated with CVD risk ^18^. These factors include nicotine use and exposure, dietary quality, physical activity, body composition, fasting blood glucose, total cholesterol, and blood pressure. Numerous studies conducted in the general population have demonstrated that higher Life’s Simple 7 scores (indicative of better CVH) are robustly associated with reduced risk of CVD-related morbidity and mortality ^19^. More recently, the Life’s Simple 7 (LS7) metric was updated to Life’s Essential 8 (LE8) and now includes sleep duration based on research indicating that insufficient (<7 hours per night) and excessive (>9 hours per night) sleep duration independently increase risk for CVD ^19^. As with LS7, data from large epidemiologic surveys suggest that better CVH as indicated by LE8 scores is associated with lower risk of CVD ^20,21^, highlighting the utility of more broadly and holistically defining CVH for identifying individuals who may benefit from earlier intervention to mitigate CVD risk.

To date, however, there is limited research examining either of these CVH frameworks in the context of pregnancy. Evidence from extant studies employing the LS7 framework indicate that the majority of individuals exhibit poor CVH in pregnancy ^22^, and that poor maternal CVH in pregnancy is associated with elevated risk of adverse obstetric outcomes (e.g., unplanned cesarean section; ^23^, greater postpartum maternal atherosclerosis ^24^ as well as worse CVH in their children ^25^. However, no study to date has explored the relationship between CVH in pregnancy and postpartum maternal psychological functioning. Therefore, it remains unclear whether variation in CVH during pregnancy relates to risk of postpartum psychological distress. Given that individuals who begin pregnancy with overweight and obesity (body mass index [BMI] ≥25kg/m^2^) are more likely to experience cardiovascular conditions during pregnancy ^26,27^ as well as postpartum psychological distress ^28,29^ compared to individuals with a BMI <25, the present study aimed to examine the relationship between changes in CVH from the second trimester of pregnancy to six months postpartum and postpartum psychological functioning among individuals who began their pregnancies with overweight or obesity. A composite score using the LE8 framework was calculated to index CVH and was comprised of body composition and CVH behaviors. CVH scores were computed with and without sleep duration included given that only a subset of participants completed the sleep assessment and sleep is a recent addition to the AHA’s current CVH framework (LE8), which has received minimal research attention in pregnancy. Therefore, a secondary aim of the present study was to evaluate whether the inclusion of sleep changed the relationship between CVH scores and postpartum psychological functioning. It was predicted that greater improvements in LS7 CVH scores would be associated with fewer symptoms of psychological distress during the postpartum period, and that the inclusion of sleep would strengthen the association between LE8 CVH scores and postpartum psychological functioning.

## Methods

### Participants and study procedures

The present study is a secondary analysis of data collected for a longitudinal study of health and well-being during the perinatal period among individuals who began their pregnancies with a BMI≥25 kg/m^2^ ^30^. Pregnant individuals (*N*=257) were recruited from local obstetrics clinics and were eligible if they had overweight or obesity prior to becoming pregnant, had a singleton pregnancy, and were ≥14 years of age at enrollment. Exclusion criteria included diagnosis of type I diabetes, taking medications or diagnosed with conditions known to influence weight, participating in a weight management program, or experiencing acute psychiatric symptoms warranting immediate intervention (e.g., suicidality). Participants 18 years and older provided written informed consent before the initiation of study procedures. Verbal assent was obtained from participants below age 18 (*n*=4) and written informed consent was provided by a parent or legal guardian. Procedures were approved by the University of Pittsburgh Institutional Review Board.

Eligible individuals attended up to seven visits over the course of the perinatal period to complete assessments of demographic (age, racial identity, marital status, educational background, household income, and parity), medical, and psychosocial factors and reported on health behavior engagement. The initial baseline assessment occurred when participants’ pregnancies were 12-20 weeks gestation (“Time 0” or T0; *n*=257). Subsequent assessments occurred at 18–22 weeks gestation (T1; *n*=253), 23–26 weeks gestation (T2: *n*=252), 27–30 weeks gestation (T3; *n*=245), 31–34 weeks gestation (T4; *n*=240), 35 weeks gestation through delivery (T5; *n*=206), and six months postpartum (T6; *n*=237). Data collection for the parent study began in September 2012 and was completed in January 2017.

### Measures of psychological distress

#### Depressive symptoms

Depressive symptoms were assessed using the Center for Epidemiologic Studies-Depression Scale (CES-D;^31^), a self-report measure of the frequency of 20 common depressive symptoms rated along a 0 (“Rarely or none of the time”) to 3 (“Most or all of the time”) Likert scale. Responses are summed to yield a total score, with higher scores reflecting more severe depressive symptoms. The CES-D has demonstrated adequate reliability and validity in a number of populations ^32–34^, including among individuals who are pregnant ^35^.

#### Perceived stress

The Perceived Stress Scale (PSS; ^36^ was administered to assess experiences of daily life stress. The PSS is a 14-item instrument on which respondents use a 0 (“never”) to 4 (“very often”) Likert scale to rate the degree to which daily life events are perceived to be uncontrollable, unpredictable, or unmanageable. Responses are summed to form a total score, with higher scores indicating more perceived stress. This scale has been shown to exhibit satisfactory reliability (Cronbach’s α = 0.85) and validity ^37^, including during the perinatal period ^38^.

### Measures of cardiovascular health

Cardiovascular health was indexed using the AHA LE8 metric ^19^, a composite score comprised of the eight health behaviors and biomarkers that are most strongly linked to individual differences in risk for cardiovascular disease. The components included in the LE8 are dietary patterns, physical activity, nicotine exposure, sleep health, BMI, blood lipids, blood glucose, and blood pressure. Of the eight components included in the LE8 composite score, the following five were evaluated as part of the study assessment battery: weight and height for calculating BMI, physical activity, nicotine use and history, dietary intake, and sleep. Data for the present study were drawn from baseline (T0) and six months postpartum (T6) visits, as these were the timepoints at which all available components of the LE8 score were measured. The measure of sleep health (described in more detail below) was added to the assessment battery in February 2015, and was therefore only available for a subset of participants. Thus, we computed a composite score which does not include sleep, reflecting domains originally in the LS7, and computed composite scores that include sleep among the participants with these data. Scoring guidelines for the LE8 were used to compute both composite scores. Complete data were available for BMI, physical activity, nicotine use and history, and dietary intake on 226 (87.9%) participants at the T0 and T6 timepoints, and of those 226 participants, sleep data were available at both timepoints on 114 (50.4%). Table 1 depicts the timepoints that each of the LE8 metrics were collected in the parent study to provide additional clarity.

**Table 1.**
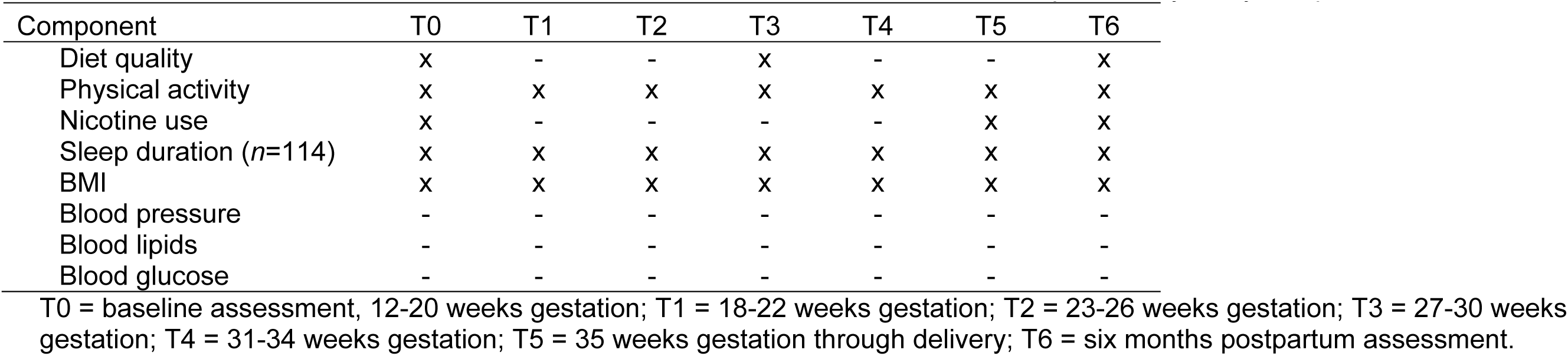
Collection of the American Heart Association’s Life’s Essential 8 metric components by study timepoint.

#### Body mass index

To calculate BMI during early pregnancy and 6-months postpartum, weight and height were objectively measured using a digital scale and a calibrated stadiometer during the T0 and T6 assessments. Participants also self-reported pre-pregnancy weight to calculate pre-pregnancy BMI.

#### Physical activity

Physical activity was assessed using the Paffenbarger Physical Activity Survey (Paffenbarger, Wing, & Hyde, 1978), a seven-day activity recall survey that evaluates the amount of physical activity due to activities of daily living (e.g., walking) and leisure activity that involves physical exertion (e.g., gardening, jogging). The Paffenbarger is a widely used instrument for assessing habitual physical activity that exhibits good reliability (Ainsworth, Jacobs, & Leon, 1993), and has been shown to correlate highly with objective measures of body composition (Choo et al., 2010) and physical activity (Prince et al., 2008). Trained staff prompted participants to recall the number of blocks walked, flights of stairs climbed, and any other physical activity for sport, exercise or recreation within the previous week. When a participant reported engaging in activity that could vary widely in intensity (e.g., using an elliptical machine), interviewers conducted follow-up questioning to obtain a more accurate assessment of intensity level (e.g., requesting distance and time). Trained raters categorized exercise and recreation activities according to the 2011 Ainsworth Compendium of Physical Activities (Ainsworth et al., 2011), a comprehensive coding system that classifies physical activities by rate of associated energy expenditure in metabolic equivalent of task (MET). Per Ainsworth, activities classified as expending 3.0-6.0 METs were considered moderate intensity PA, while activities classified as expending ≥6.0 METs were considered to be in the vigorous intensity range (Ainsworth et al., 2011). All surveys were rated by two individuals, the interviewer who administered the survey and a second independent reviewer. In the case of a discrepant classification, the activity with the lower MET value was chosen, as this was considered to be a more conservative approach. MET values associated with each activity were then used to calculate the number of minutes spent engaging in moderate or vigorous physical activity. In accordance with the AHA LE8 guidelines for scoring physical activity, minutes of vigorous PA were multiplied by two. Minutes of moderate or vigorous physical activity were then summed to obtain total minutes per week.

#### Nicotine use and history

Participants provided information on their current nicotine use, and if applicable, years of nicotine use, age of onset, number of quit attempts, and quit date, on a health survey. For participants who formerly used nicotine, quit date was subtracted from the date of assessment to determine number of years quit, which was then used to derive the nicotine use score. The health survey was updated shortly after data collection began to include additional questions on other nicotine delivery systems (NDS) apart from combustible tobacco (e.g., e-cigarettes, vaporizers, hookah). Therefore, while all participants provided information about use of combustible tobacco, 15 participants were missing data on use of other NDS due to having completed the survey prior to the revision. Given evidence suggesting that the prevalence of NDS use compared to combustible tobacco use was far lower during the period in which data was collected for the parent study ^39^, data for the 15 participants who were missing information on NDS use were retained for the calculation of LE8 nicotine use scores.

#### Dietary intake and quality

At T0 and T6 timepoints, participants completed two 24- hour dietary recall interviews either over the phone or in person during the baseline assessment. Dietary recall interviews were conducted to obtain a detailed record of all foods and beverages consumed in the previous 24 hours and designed to capture one weekday and one weekend day given evidence that reports of dietary intake vary significantly depending on what point during a week the recall occurs ^40^. Interviews were administered by master’s-level clinicians who received certifications after completing on-site training in Nutrition Data System for Research software, Nutrition Coordinating Center Food and Nutrient Database, conducting dietary interviews, and dietary recall quality assurance (University of Minnesota, 2023). The Nutrition Data System for Research ^41^ analysis software was then used to calculate the 2015 version of the Healthy Eating Index (HEI) based on the dietary intake data obtained from the two food recall interviews, from which an average HEI score was derived. The HEI is a measure of dietary quality developed to quantify the degree to which an individual’s dietary intake patterns conform to the recommendations put forth in the 2015-2020 Dietary Guidelines for Americans ^42^. The HEI is comprised of 13 components, and scores on each component are summed to form a total HEI score ranging from 0-100, with higher scores indicating dietary intake more closely aligned with dietary guidelines. In the general population, higher HEI-2015 scores have been associated with lower all-cause mortality, and reduced risk of mortality specifically from cardiovascular disease, Type II Diabetes, and cancer ^43–45^, and the HEI-2015 has been shown to exhibit satisfactory construct and criterion validity ^44^. Data on the psychometric properties of the HEI-2015 during the perinatal period are limited. However, HEI-2015 scores have previously been used in the assessment of CVH during pregnancy ^22^.

#### Sleep duration

Participants completed the Pittsburgh Sleep Quality Index (PSQI; ^46^, a 19-item self-report measure on which respondents rate their sleep patterns in the past month using a 0 to 3 Likert Scale. The PSQI has been validated for use in a variety of populations ^47^, including among individuals who are pregnant ^48,49^. For the purposes of the present study, total sleep duration as reported on the PSQI was used to calculate the sleep component of the LE8 score.

#### Life’s Essential 8 scoring

Scoring guidelines for the AHA’s LE8 metric have been updated from the LS7 formulation to increase sensitivity for detecting individual differences in CVH. Previously, individual components were rated as “ideal”, “intermediate”, or “poor” based on a set of pre-defined criteria, which obscured the impact of within-component variation on outcomes of interest. The revised scoring algorithm for the LE8 now assigns each individual component a score ranging from 0-100 points, which are used to create a global CVH metric by calculating the unweighted average of all included component scores. The criteria for scoring each individual component were defined by the AHA LE8 working group ^19^ and are described in Table 2. The AHA has developed different scoring systems for adults (i.e., age ≥20) and children (i.e., age ≤19) to account for developmental factors that impact CVH metrics ^19^. Nevertheless, participants aged <20 in the current study (*n*=15; age range=15.39-19.92 years) were scored as adults based on evidence that diet quality, physical activity, nicotine use, and BMI measured during pregnancy do not differ significantly by age when comparing adults to late adolescents ^39,50–53^. As previously noted, both because sleep is a new addition to the AHA’s composite CVH score and because significantly more participants had complete data on BMI, PA, nicotine use and history, and dietary intake at both timepoints than did on sleep, LE8 scores were calculated both with and without sleep. To evaluate the impact of longitudinal change in CVH from T0 to T6, a CVH change score was calculated by subtracting LE8 scores at T0 from LE8 scores at T6. This was done for LE8 scores with and without sleep included, which permitted comparisons of the effect of CVH scores without sleep to the effect of CVH scores with sleep.

**Table 2.**
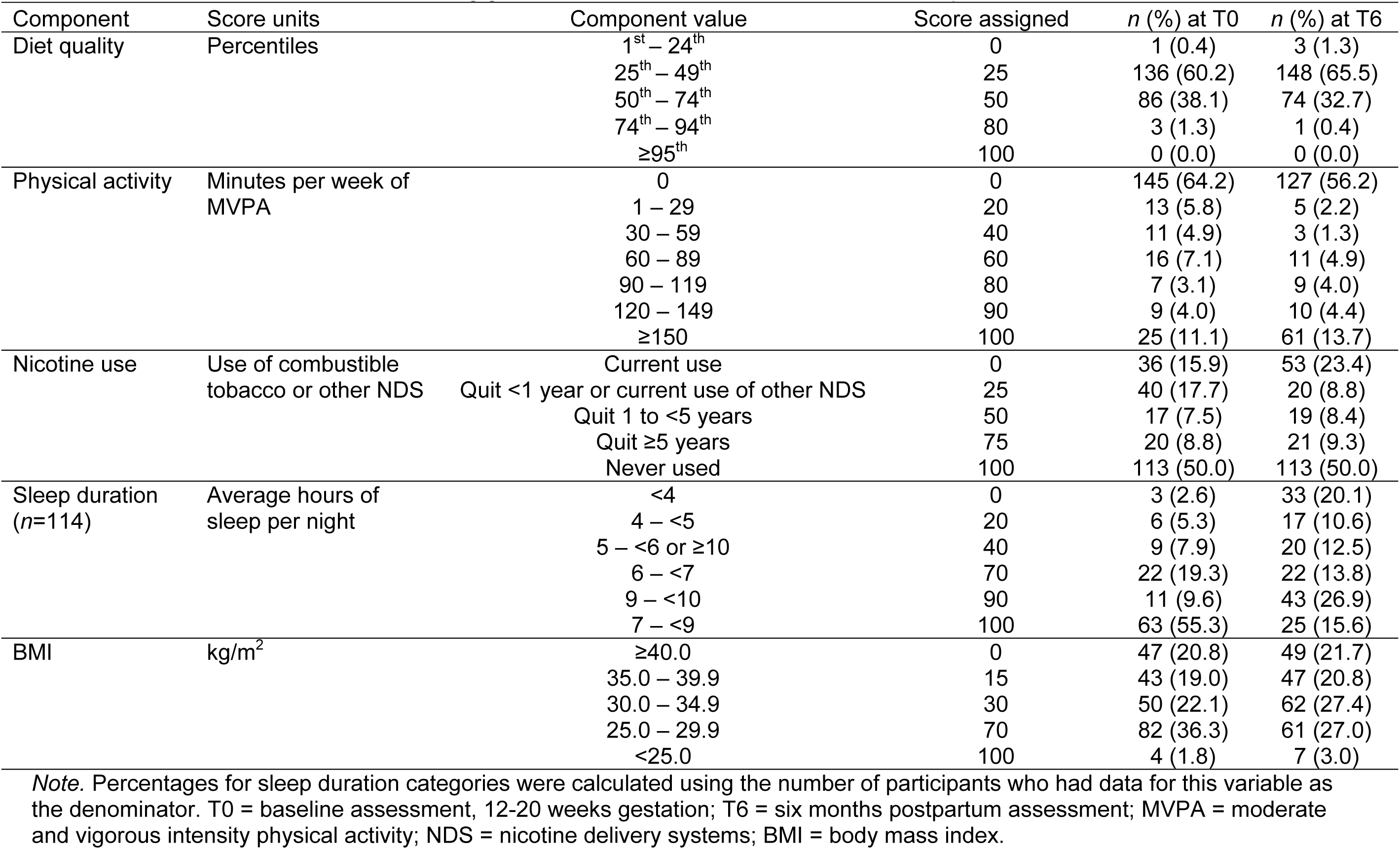
American Heart Association scoring guidelines for the Life’s Essential 8 metric components.

#### Statistical Approach

Prior to hypothesis testing, all data were examined to assess missingness, identify extreme values, and confirm that the data structure met analytic assumptions (e.g., normality). A square root transformation was applied to CES-D scores to adjust for positive skew. Univariate outlier detection was conducted using Rosner’s Test ^54^ in the R package EnvStats ^55^. Descriptive analyses were conducted to examine sample characteristics. Multiple linear regression analyses were then performed to evaluate whether changes in LE8 scores from T0 to T6 were associated with T6 CES-D and PSS scores. Analyses were performed with and without sleep included in the LE8 metric. All models were adjusted for baseline LE8 scores to account for the influence of individual variation on CVH during the second trimester of pregnancy on change in CVH through six months postpartum. In addition, given evidence that health behavior engagement changes over the course of pregnancy ^56–59^ and that demographic characteristics and social determinants of health are associated with differences in CVD risk ^19,60^, weeks gestation of pregnancy at the time of enrollment, age, racial identity, household income (dichotomized as ≤$30,000 or >$30,000 per year), and educational attainment were included as covariates in all models. Baseline scores on the CES-D and PSS were also included as covariates in the relevant models. Model fit was evaluated using the overall F-test and regression diagnostic plots were visually inspected to confirm that the assumptions of linear regression were met. Presence of high leverage outliers was evaluated via Cook’s distance values using a cutoff of ≥ 0.5; no values exceeded this threshold for any analysis. Post-hoc paired-sample t-tests were performed to evaluate differences between baseline and postpartum LE8 component scores. For all tests, the level of statistical significance was set at *p*<0.05 and standardized coefficients were selected for reporting significant effects. Analyses were conducted in R Studio ^61^ using R version 4.2.2 ^62^.

## Results

### Sample characteristics

Participants completed their initial baseline assessment visit when their pregnancies were 15.64 (*SD*=2.45) weeks gestation. Mean CVH scores excluding sleep were 40.27 (*SD*=17.64) at baseline and 41.97 (*SD*=19.98) at six months postpartum. With the inclusion of sleep in the CVH metric, mean CVH scores were 55.05 (*SD*=15.03) at baseline and 46.86 (*SD*=17.92) at six months postpartum. Mean postpartum depressive symptoms were in the mild range (*M*=10.75, *SD*=9.58, range=0-49), and 51 individuals (22.6%) scored above the clinical cutoff of 16 suggestive of risk for a depressive episode. Ratings of perceived stress were in the moderate range of severity (*M*=20.73, *SD*=8.97, range=3-45). Demographic and clinical characteristics of the sample at each timepoint are presented in more detail in Table 3.

**Table 3.** Demographic and clinical characteristics of sample to date.

|  | T0 | T6 |
| --- | --- | --- |
|  | Mean (SD) | Mean (SD) |
| Weeks gestation | 15.64 (2.45) | - |
| Pre-pregnancy BMI | 32.74 (6.55) | - |
| Age (years) | 28.43 (5.40) | - |
| CES-D | 12.46 (9.88) | 10.75 (9.58) |
| PSS | 20.91 (8.73) | 20.73 (8.97) |
| CVH total (sleep included; *n*=114) | 53.05 (15.03) | 46.86 (17.92) |
| CVH total (sleep excluded) | 40.27 (17.64) | 41.97 (19.98) |
| BMI | 34.17 (7.15) | 34.94 (7.41) |
| 2015 HEI Scores<sup>a</sup> | 47.36 (10.96) | 45.73 (10.78) |
| Minutes of weekly MVPA<sup>b</sup> | 120.15 (293.96) | 138.01 (316.50) |
| Hours of sleep per night (*n*=114) | 7.05 (1.62) | 6.12 (3.22) |
|  | *n* (%) | *n* (%) |
| CES-D scores $\geq 16^c$ | 65 (28.76) | 51 (22.56) |
| Current combustible tobacco or other NDS use | 52 (23.01) | 72 (31.85) |
| Yearly household income |  | - |
| ≤ \$30,000 | 148 (65.48) | |
| $>$ \$30,000 | 78 (34.51) | |
| Education |  | - |
| Grade school or some high school | 28 (12.39) |  |
| High school graduate or GED | 47 (20.80) |  |
| Some college or technical school | 86 (38.05) |  |
| 4-year college graduate | 31 (13.72) |  |
| Postgraduate degree | 34 (15.04) |  |
| Racial Identity |  | - |
| Asian | 1 (0.44) |  |
| American Indian or Alaska Native | 3 (1.32) |  |
| Black or African American | 113 (50.0) |  |
| Multi-racial | 9 (3.98) |  |
| Unknown | 4 (1.77) |  |
| White | 96 (42.48) |  |
| Hispanic ethnicity | 7 (6.8) | - |

*Note.* T0 = baseline assessment; T6 = postpartum assessment; BMI = body mass index; CES-D = Center for Epidemiologic Studies Depression Scale; PSS = Perceived Stress Scale; CVH = cardiovascular health; HEI = healthy eating index; MVPA = moderate and vigorous intensity physical activity; NDS = nicotine delivery systems; GED = General Educational Development.
<sup>a</sup> HEI scores for the weekend and weekday dietary intake assessments were averaged to create a single HEI score representing overall diet quality during the week of assessment.
<sup>b</sup> Minutes of vigorous physical activity were not doubled when calculating descriptive statistics presented in this table. They were only doubled for the purposes of calculating the Life's Essential 8 metric, per the American Heart Association's scoring guidelines.
<sup>c</sup> Scores of 16 or more on the CES-D are considered indicative of risk for a major depressive episode.

### Comparison of individuals with and without sleep data

As described above, because the PSQI was added to the assessment battery midway through the study, only 114 participants (50.4%) completed it at the baseline assessment. At baseline, there were significant differences between those with and without PSQI data in terms of household income (*X*^2^(1, *N*=226) = 6.56, *p*=0.01) and weeks gestation of pregnancy at the time of enrollment (*β*=-0.18, *p*<0.01). Regarding household income, 57% of individuals who completed the PSQI reported earning ≤$30,000 compared to 74% of individuals who did not complete the PSQI. Individuals who completed the PSQI also entered the study later in their pregnancies compared to those who did not complete the PSQI (Mean weeks gestation with vs. without PSQI: 16.67±2.39 vs. 14.65±2.09). There were no other significant differences in baseline demographic characteristics between those with and without PSQI data (*p*s>0.06). When comparing those with and without PSQI data on postpartum outcomes, individuals who completed the PSQI exhibited significantly higher CVH scores calculated excluding sleep (*M*=44.77, *SD*=20.31) compared to those who did not complete the PSQI (*M*=39.12, *SD*=19.31; *β*=-0.18, *p*<0.01). There were no significant differences between those with and without PSQI data on postpartum CES-D or PSS scores (*p*s>0.36).

### Changes in individual CVH components from baseline to postpartum

Compared to baseline, participants had significantly lower scores for BMI (*t*(225)=2.92, *p*<0.01) and sleep (*t*(112)=5.69, *p*<0.01), and significantly higher scores for PA (*t*(225)=-3.91, *p*<0.01) at the postpartum assessment. There were no significant differences from baseline to postpartum in LE8 component scores for dietary intake or nicotine use (*p*s>0.06).

### Relationship between CVH behaviors and postpartum psychological distress

When excluding sleep from the CVH metric, worsening CVH scores from baseline to six months postpartum predicted higher postpartum depressive symptoms (*β*=-0.18, *p*<0.01) and ratings of perceived stress (*β*=-0.13, *p*=0.02). When examining unadjusted means, compared to those whose CVH improved by >1SD, individuals whose CVH worsened by >1SD scored 6.42 points higher on the CES-D (*M*CES-D=15.25±10.92 vs. 8.52±6.90) and 6.12 points higher on the PSS (*M*PSS=24.45±8.29 vs. 17.83±8.70) at six-month postpartum. Adjusting for covariates, this difference in mean scores persisted, with individuals whose CVH scores worsened by >1SD reporting higher severity symptoms compared to those whose scores improved by >1SD (*M*CES-D=3.53±0.78 vs. 2.59±0.81; *M*PSS=23.34±8.97 vs. 17.88±5.69), though the difference was attenuated. The impact of adjustment on mean scores was more pronounced for the CES-D than the PSS, which is consistent with results from regression models demonstrating that covariates were more strongly associated with CES-D scores than they were PSS scores. Further, improved CVH was associated with lower odds of having depressive symptom severity above the cutoff score of 16 typically used to identify the presence of a depressive episode (odds ratio=0.976, *p*=0.48). These relationships persisted after adjusting for baseline CVH, baseline symptom scores, as well as demographic factors such as age, racial identity, educational attainment, and household income. Figure 1 depicts differences between postpartum CES-D and PSS scores for those whose CVH improved vs. worsened from pregnancy to postpartum. Table 4 includes more detailed results from linear regression models using the CVH metric that excluded sleep.

**Figure 1.**
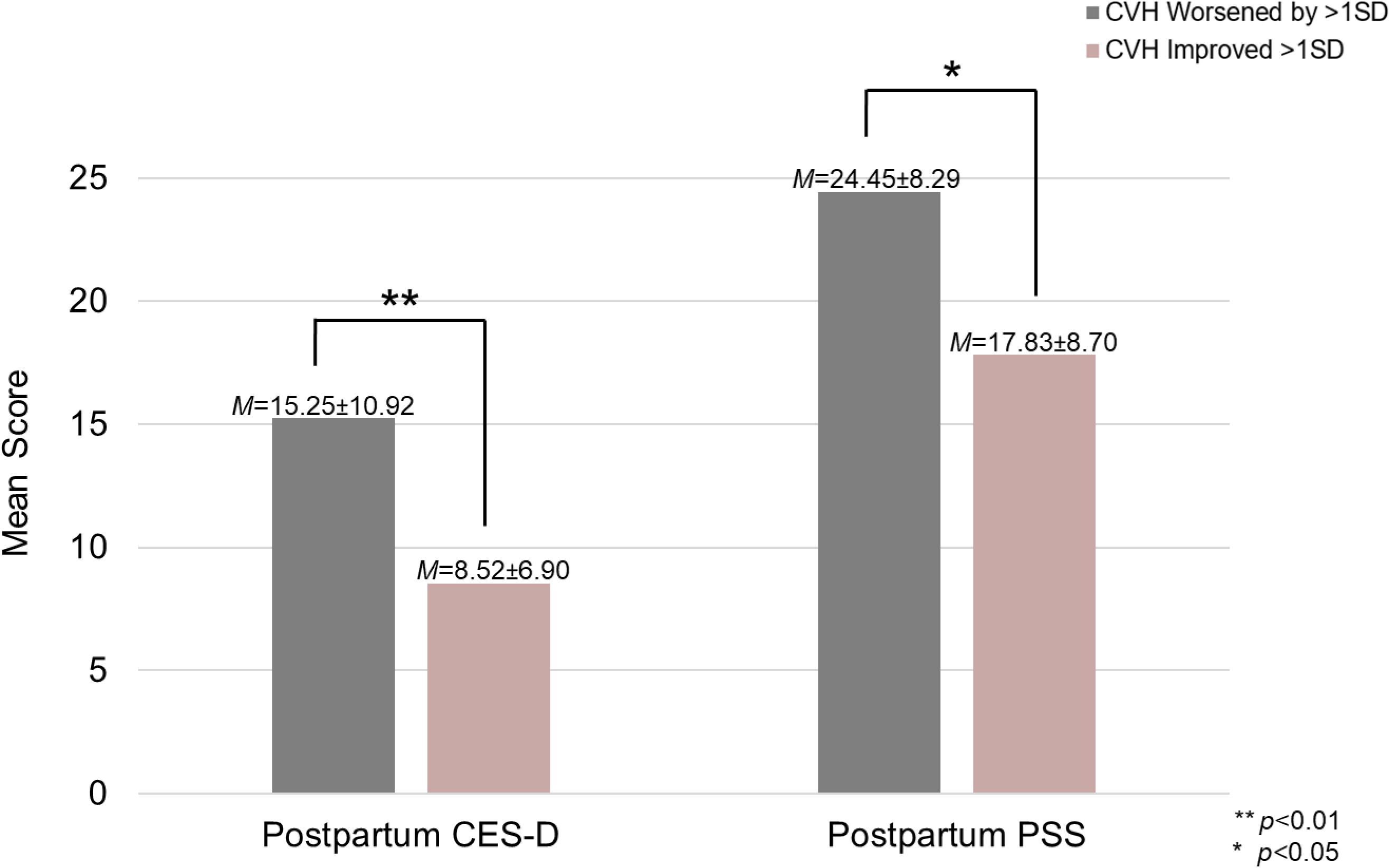
Differences in postpartum CES-D and PSS scores among participants whose CVH worsened from pregnancy to postpartum compared to those whose CVH scores improved. *Note*. CES-D = Center for Epidemiologic Studies Depression Scale; PSS = Perceived Stress Scale; CVH = cardiovascular health

**Table 4.** Associations between the changes in CVH (excluding sleep) from baseline to six-months postpartum and postpartum psychological functioning.

| Coefficient | Estimate (SE) | p |
| --- | --- | --- |
| **Model: CES-D** |  |  |
| (Intercept) | **2.076 (0.624)** | **0.008** |
| **Change in CVH scores from T0 to T6** | **-0.017 (0.005)** | **0.002** |
| T0 CVH scores | 0.002 (0.006) | 0.679 |
| **T0 CES-D scores** | **0.071 (0.009)** | **<0.001** |
| T0 Age | -0.002 (0.019) | 0.570 |
| T0 Gestational age in weeks | -0.020 (0.035) | 0.911 |
| **Black racial identity** | **0.453 (0.225)** | **0.045** |
| Asian racial identity | 0.043 (1.261) | 0.973 |
| American Indian or Alaska Native racial identity | -0.074 (0.748) | 0.921 |
| Multi-racial identity | 0.479 (0.449) | 0.287 |
| Unknown racial identity | 1.104 (0.754) | 0.286 |
| High school graduate/GED | -0.249 (0.296) | 0.399 |
| Some college or technical school | -0.382 (0.282) | 0.177 |
| Four-year college degree | -0.593 (0.403) | 0.183 |
| **Postgraduate degree** | **-0.906 (0.442)** | **0.006** |
| Annual household income > \$30,000 | 0.399 (0.305) | 0.191 |
| Coefficient | Estimate (SE) | p |
| **Model: PSS** |  |  |
| (Intercept) | 2.179 (5.138) | 0.672 |
| **Change in CVH scores from T0 to T6** | **-0.078 (0.032)** | **0.018** |
| T0 CVH scores | 0.047 (0.035) | 0.181 |
| **T0 PSS scores** | **0.607 (0.059)** | **<0.001** |
| T0 Age | 0.133 (0.204) | 0.513 |
| T0 Gestational age in weeks | 0.134 (0.108) | 0.217 |
| Black racial identity | 0.296 (1.293) | 0.819 |
| Asian racial identity | 2.715 (7.209) | 0.707 |
| American Indian or Alaska Native racial identity | -1.787 (4.265) | 0.676 |
| Multi-racial identity | -0.739 (2.571) | 0.774 |
| Unknown racial identity | -3.177 (4.329) | 0.464 |
| High school graduate/GED | -1.616 (1.692) | 0.341 |
| Some college or technical school | -2.007 (1.601) | 0.211 |
| Four-year college degree | -3.096 (2.309) | 0.182 |
| **Postgraduate degree** | **-5.063 (2.521)** | **0.046** |
| Annual household income > \$30,000 | 0.789 (1.732) | 0.649 |

*Note.* Bolded p-values indicate statistical significance ( $p < 0.05$ ). Reference groups for categorical variables are as follows: White racial identity; less than a high school education or equivalent; annual household income ≤ \$30,000. T0 = baseline assessment; T6 = postpartum assessment; CES-D = Center for Epidemiologic Studies Depression Scale; PSS = Perceived Stress Scale; CVH = cardiovascular health; GED = General Educational Development.

### Relationship between CVH behaviors, including sleep, and postpartum psychological distress

When sleep was included in the CVH metric, the associations between change in CVH scores and postpartum symptom scores were no longer significant (CES-D model: *β*=0.06, *p*=0.4; PSS model: *β*=0.04, *p*=0.6). Post-hoc assessment of the correlation between sleep duration early in pregnancy and postpartum symptom scores indicated that sleep duration at T0 was weakly negatively associated with T6 depressive symptoms (*r*=-0.09, *p*=0.30) and not correlated with T6 perceived stress (*r*=-0.001, *p*=0.99). In contrast, sleep measured at T6 was strongly positively correlated with concurrent depressive symptoms (*r*=0.40, *p*<0.01) and perceived stress (*r*=0.31, *p*<0.01), meaning that longer postpartum sleep duration was associated with higher severity symptomatology. Table 5 provides more detailed results from linear regression models using the CVH metric that included sleep.

**Table 5.** Associations between the changes in CVH including sleep from baseline to six-months postpartum and postpartum psychological functioning. (*N=*114)

| Coefficient | Estimate (SE) | *p* |
| --- | --- | --- |
| **Model: CES-D** |  |  |
| **(Intercept)** | **4.363 (1.574)** | **0.007** |
| Change in CVH scores from T0 to T6 | 0.005 (0.008) | 0.493 |
| T0 CVH scores | -0.001 (0.010) | 0.957 |
| **T0 CES-D scores** | **0.077 (0.014)** | **<0.001** |
| T0 Age | -0.051 (0.027) | 0.067 |
| T0 Gestational age in weeks | -0.041 (0.052) | 0.434 |
| Black racial identity | 0.607 (0.332) | 0.071 |
| Asian racial identity | 0.673 (1.272) | 0.598 |
| American Indian or Alaska Native racial identity | -0.161 (1.328) | 0.903 |
| Multi-racial identity | 1.516 (0.961) | 0.118 |
| Unknown racial identity | 1.395 (0.787) | 0.079 |
| High school graduate/GED | -0.886 (0.511) | 0.086 |
| Some college or technical school | -0.668 (0.479) | 0.166 |
| Four-year college degree | -1.057 (0.604) | 0.083 |
| **Postgraduate degree** | **-1.577 (0.665)** | **0.020** |
| **Annual household income > \$30,000** | **1.046 (0.423)** | **0.015** |
| Coefficient | Estimate (SE) | *p* |
| **Model: PSS** |  |  |
| **(Intercept)** | **11.702 (9.599)** | **0.226** |
| Change in CVH scores from T0 to T6 | 0.025 (0.048) | 0.606 |
| T0 CVH scores | 0.019 (0.062) | 0.763 |
| **T0 PSS scores** | **0.604 (0.085)** | **<0.001** |
| T0 Age | -0.132 (0.167) | 0.427 |
| T0 Gestational age in weeks | 0.056 (0.318) | 0.860 |
| Black racial identity | 1.103 (2.008) | 0.584 |
| Asian racial identity | 5.634 (7.669) | 0.486 |
| American Indian or Alaska Native racial identity | -0.369 (8.055) | 0.964 |
| Multi-racial identity | 1.018 (5.776) | 0.783 |
| Unknown racial identity | -1.315 (4.765) | 0.783 |
| High school graduate/GED | -3.173 (3.050) | 0.301 |
| Some college or technical school | -2.256 (2.856) | 0.432 |
| Four-year college degree | -3.271 (3.672) | 0.375 |
| Postgraduate degree | -5.816 (4.002) | 0.149 |
| Annual household income > \$30,000 | 4.013 (2.533) | 0.116 |

*Note.* Bolded p-values indicate statistical significance ( $p < 0.05$ ). Reference groups for categorical variables are as follows: White racial identity; less than a high school education or equivalent; annual household income ≤ \$30,000. T0 = baseline assessment; T6 = postpartum assessment; CES-D = Center for Epidemiologic Studies Depression Scale; PSS = Perceived Stress Scale; CVH = cardiovascular health; GED = General Educational Development.

## Discussion

The present study examined the longitudinal association between change in CVH from early pregnancy to the postpartum period and postpartum psychological distress in a community sample of individuals with BMI ≥ 25. The AHA’s Life’s Essential 8 composite metric was used to index CVH, calculated both with and without sleep – a new addition to the AHA’s composite metric - to capitalize on the larger sample of individuals with available data on BMI, nicotine exposure, PA, and diet quality but who did not complete the sleep assessment. Consistent with study hypotheses, improvements in CVH from pregnancy to six months postpartum was associated with lower severity of depressive symptoms and perceived stress when excluding sleep from the CVH metric, relationships which persisted after adjusting for the potentially confounding effects of early pregnancy sociodemographic characteristics, CVH, and symptom measures. To our knowledge, this is the first study to examine the relationship between CVH during pregnancy as measured using the AHA’s LE8 composite and postpartum psychological outcomes. Prior research exploring how cardiovascular conditions emerging during pregnancy relate to postpartum psychological functioning have predominantly focused on the impact of diagnosed cardiovascular illness. Thus, our findings extend the existing evidence by demonstrating that cardiovascular health when measured more holistically than the presence or absence of diagnosable disease is associated with postpartum psychological health postpartum.

These findings suggest that the AHA’s measure of CVH may be useful for identifying individuals prior to delivery who are at risk for experiencing postpartum depression and elevated stress. Importantly, the factors that are included in the Life’s Essential 8 metric are either routinely collected throughout pregnancy (i.e., weight, blood biomarkers) or are relatively convenient to assess using self-report measures. Therefore, the potential impact to current clinical workflows would be fairly minor, especially when considered in relation to the benefits of monitoring CVH during pregnancy. For example, using this metric for early identification of vulnerable individuals will enable providers to connect patients to prevention and intervention resources to optimize postpartum health and well-being. Given the pernicious effects of postpartum mental health conditions such as depression on maternal suicide risk ^63^ and infant development ^64^, improving screening and identification of pregnant individuals who are vulnerable to postpartum distress using the CVH metric has the potential to engender widespread benefits to maternal and child health.

Another potential implication of this observed link between change in CVH over pregnancy and postpartum psychological distress is that interventions targeting CVH during pregnancy may improve maternal well-being following delivery. This idea is consistent with evidence that participating in structured physical activity during pregnancy reduces risk of postpartum depression ^65^. An emerging body of research has also demonstrated that brief interventions for insomnia during the perinatal period may reduce risk for postpartum depression ^66,67^, though this area of research is relatively nascent. It will be important to conduct additional research exploring whether interventions focused on aspects of CVH yield similar benefits to postpartum mental health.

Contrary to our hypotheses, CVH was no longer associated with postpartum psychological distress when sleep was included as a component of CVH. That the addition of sleep to the CVH metric changed the link to psychological distress stands in contrast to previous research demonstrating that poor sleep quality during the perinatal period is associated with increased risk of experiencing postpartum symptoms such as depression and anxiety ^68–71^. However, given that only 50% of the present sample (*n*=114) completed the sleep assessment at baseline, the lack of an association between change in CVH and postpartum psychological distress when including sleep may be attributable to the significant sample loss incurred when by doing so. Further, there were significant differences in household income, weeks gestation of pregnancy at the time of enrollment, and postpartum CVH (excluding sleep) between those did not have sleep data (i.e., were recruited earlier in the study period) and those who did, suggesting possible cohort effects. It is also important to note that sleep duration early in pregnancy was only weakly correlated with postpartum depressive symptoms and not correlated with postpartum ratings of perceived stress in the present study. Therefore, it is possible that the lack of an association between change in CVH and postpartum psychological distress when including sleep duration as a component of the CVH metric may be explained by the fact that early pregnancy sleep duration was not robustly related to postpartum psychological symptom scores in this sample. Further, sleep measured at six months postpartum was strongly positively correlated with concurrent depressive symptoms and perceived stress, suggesting that individuals whose postpartum sleep duration was in the more ideal range were experiencing higher severity symptoms. This pattern is inconsistent with prior research examining the relationship between sleep duration and postpartum psychological functioning ^68,72,73^. Finally, sleep duration is only one of several sleep characteristics that has been linked to health and well-being outcomes (e.g., sleep discontinuity, time spent awake after initiating sleep), and it is unknown whether sleep duration is the characteristic that is most salient during pregnancy and postpartum. Of note, sleep duration recommendations differ by age group for calculating LE8 among children, and it is possible that adjustments to LE8 scoring guidelines for sleep may similarly be warranted during pregnancy. Given that sleep disruption is a common occurrence during the perinatal period, it is possible that the relationship between sleep and postpartum psychological functioning manifests differently during this period. Additional research is needed to better understand how sleep difficulties that are common during the perinatal period impact postpartum health and psychological well-being.

### Strengths, limitations, and directions for further research

In addition to being the first study to employ the AHA’s Life’s Essential 8 framework for assessing the relationship between CVH and psychological functioning during the perinatal period, there are other notable strengths of the present study to highlight. We focused our investigation on individuals who began their pregnancies with overweight or obesity based on evidence that this is a population at heightened risk for cardiovascular conditions acquired in pregnancy, obstetric complications, and poor postpartum mental health ^27,28^. As such, it is important to understand the relationship between indicators of CVH and psychological functioning in this vulnerable group to permit more effective prevention and intervention efforts. In addition, a substantial proportion of the sample self-identified as being Black or African American (50%) and the majority reported a yearly household income of $30,000 or less (65%). Prior research, in contrast, has primarily been conducted in samples that are predominantly White identifying and higher income, potentially hampering efforts to better understand the significant inequities in perinatal health and well-being for individuals from disadvantaged and marginalized communities ^74^.

Despite these strengths, there are a number of important limitations that should be taken into account when interpreting our findings. First, sleep data were only available for half of the sample, restricting statistical power to examine the relationship between CVH and postpartum psychological functioning using the full set of CVH components measured in the parent study. Further, because we did not collect blood samples or measure blood pressure, CVH scores were limited to BMI and health behaviors known to predict CVD risk. Therefore, we were unable to comprehensively assess the impact of CVH as conceptualized by the AHA on postpartum psychological outcomes. Second, given the dearth of research employing the Life’s Essential 8 metric in the perinatal period, it is unclear whether it is necessary to adapt the metric to account for the unique context of pregnancy. For example, pregnancy is associated with normative changes in health behaviors such as diet (e.g., dietary restrictions, vitamin supplementation), weight, and blood pressure that may or may not be relevant for estimating disease risk. Indeed, it is not yet established whether the factors currently included in the CVH metric are the strongest predictors of cardiovascular health or CVD risk in childbearing individuals, given that the metric was developed based on research conducted in the general population without regard for the impact of pregnancy. Relatedly, it may be important to consider whether other measures of body weight and composition should be added to the metric beyond BMI, such as gestational weight gain and postpartum weight retention. Additional research exploring these questions is necessary to determine whether the LE8 metric as it is currently composed and scored is appropriate for evaluating CVH during the perinatal period. Finally, sleep and physical activity were assessed using self-report, which has been shown to correlate only moderately with objective measures of these behaviors ^75,76^. Future studies evaluating CVH in the perinatal period would benefit from employing actigraphy to obtain more robust, accurate, and nuanced measures of these behaviors.

## Conclusions

The present study demonstrated that worsening of cardiovascular health behaviors from pregnancy to postpartum is longitudinally associated with more severe depressive symptoms and greater perceived stress at 6-months postpartum among individuals at high risk for future CVD. These findings provide initial evidence that improved management of cardiovascular risk factors during pregnancy may confer specific benefits to maternal mental health in addition to reducing the likelihood of developing pregnancy-related cardiovascular conditions. Additional research with more robust and complete measurement of the components of CVH across the perinatal period is needed to further validate these associations, and to explore whether interventions targeting CVH may promote maternal mental health.

## Data Availability

De-identified data can be provided upon reasonable request.

